# Virtual Reality as a Non-Pharmacological Intervention for Pediatric Preoperative Anxiety: A Quality Improvement Initiative and Retrospective Analysis

**DOI:** 10.64898/2026.01.21.26344597

**Authors:** Walaa S. Abu Rukbah, Narendra Vadlamudi, Siri Garrett, Suad Alshammari, Dayanjan S Wijesinghe

**Affiliations:** Department of Pharmacy Practice, Faculty of Pharmacy, University of Tabuk, Tabuk, Saudi Arabia; Children’s Hospital of Richmond at Virginia Commonwealth University, Richmond, VA 23219; Department of Clinical Practice, College of Pharmacy, Northern Border University, Rafha, Saudi Arabia; Department of Pharmacotherapy and Outcome Sciences, School of Pharmacy, Virginia Commonwealth University, Richmond VA 23298

## Abstract

Preoperative anxiety affects 50-75% of pediatric surgical patients and can lead to adverse perioperative outcomes including difficult anesthesia induction, emergence delirium, and prolonged recovery. Virtual reality has emerged as a promising non-pharmacological intervention, yet evidence from real-world clinical implementation remains limited. We conducted a nine-month quality improvement initiative at VCU Children’s Hospital from October 2023 through June 2024, implementing custom-designed virtual reality interventions for pediatric patients aged 6-15 years undergoing elective surgical procedures. The intervention was refined through five iterative Plan-Do-Study-Act cycles before retrospective analysis of prospectively collected data from 80 pediatric patients, 65 parents or caregivers, and 29 preoperative nurses. Primary outcomes included distress measured via 0-10 Numerical Rating Scale and fear assessed using the 0-4 Children’s Fear Scale, both measured pre- and post-intervention. Secondary outcomes included physiological parameters, parent anxiety, and stakeholder satisfaction. VR intervention demonstrated significant reductions in both patient distress (median change: -2.0 points, 95% CI [-2.00, -1.00], p<0.001) and fear (median change: -1.0 point, 95% CI [-1.00, 0.00], p<0.001). A majority of patients achieved a clinically significant reduction (≥30%) in both distress (66.2%) and fear (52.5%). Physiological markers demonstrated significant reductions in heart rate (mean decrease 5.8 beats per minute, p<0.001) and systolic blood pressure (mean decrease 4.1 mmHg, p<0.0001). Baseline anxiety levels predicted treatment response, with higher initial scores associated with greater likelihood of improvement. Stakeholder satisfaction was consistently high across patients (91.2% willing to use virtual reality again), parents (95.4% would recommend), and nurses (100% endorsement, zero workflow disruption). Virtual reality intervention significantly reduces pediatric preoperative anxiety with high stakeholder acceptance and minimal workflow disruption, supporting integration into routine preoperative care protocols.

**AUTHOR SUMMARY:** We developed and implemented a custom virtual reality intervention to reduce anxiety in children before surgery. Preoperative anxiety is extremely common in pediatric patients and can negatively affect their surgical experience and recovery. Over nine months, our team created child-friendly virtual reality environments and systematically integrated this technology into routine care at a busy pediatric surgical unit, refining our approach based on continuous feedback from patients, families, and healthcare providers.

Our findings demonstrate that children using virtual reality showed substantial reductions in both self-reported anxiety and objective physiological measures like heart rate and blood pressure. Importantly, children who were most anxious before surgery experienced the greatest benefit from the intervention. Nearly all children expressed willingness to use virtual reality again in future procedures, and parents overwhelmingly recommended it for other families facing similar situations. Nursing staff found the technology easy to incorporate into their workflow without causing procedural delays or disrupting standard care.

This work demonstrates that virtual reality can be successfully implemented in real-world clinical settings as a safe, effective, non-pharmacological approach to managing surgical anxiety in children. By reducing fear and distress during one of the most challenging moments in a child’s healthcare journey, virtual reality has significant potential to improve the preoperative experience for pediatric patients and their families.

## INTRODUCTION

Preoperative anxiety represents one of the most prevalent and clinically significant challenges in pediatric surgical care, affecting 50-75% of children undergoing surgical procedures(1,2). This anxiety manifests through psychological distress, physiological stress responses, and behavioral difficulties that can compromise both immediate perioperative safety and long-term psychological outcomes(3,4). Children experiencing high preoperative anxiety demonstrate increased risk of difficult anesthesia induction, emergence delirium, postoperative pain, prolonged recovery times, and development of negative healthcare attitudes that persist into subsequent medical encounters (1).

Current approaches to managing pediatric preoperative anxiety include pharmacological premedication, behavioral preparation programs, and parental presence during induction. While oral midazolam remains the most commonly employed anxiolytic, it carries risks of paradoxical reactions, respiratory depression, and delayed discharge (2,3,5). Non-pharmacological interventions such as therapeutic play, music therapy, and clown therapy have demonstrated anxiety-reducing effects but require specialized personnel and may not be universally available or acceptable to all children(2,6). These limitations have driven investigation of novel technological interventions that can provide consistent, accessible, and effective anxiety management without pharmacological side effects.

Virtual reality technology has emerged as a promising non-pharmacological intervention for procedural anxiety and pain management across diverse medical contexts. The theoretical foundation for virtual reality’s anxiolytic effects derives from attention-based distraction models, which posit that immersive virtual environments engage cognitive and sensory resources sufficiently to reduce processing of anxiety-provoking stimuli(4,7). By creating compelling alternative realities that capture visual, auditory, and proprioceptive attention, virtual reality may interrupt the anxiety response cycle and reduce both subjective distress and objective physiological arousal.

Despite growing interest in pediatric virtual reality applications, significant knowledge gaps persist regarding real-world clinical implementation. Most existing studies have examined virtual reality in controlled research settings with brief procedural applications, leaving questions about feasibility, stakeholder acceptance, and sustainable integration into routine clinical workflows(1,3,8,9). Furthermore, few investigations have employed comprehensive outcome assessment encompassing psychological, physiological, and experiential measures across multiple stakeholder groups including patients, families, and healthcare providers.

To address these gaps, we conducted a nine-month quality improvement initiative implementing custom-designed virtual reality interventions for pediatric preoperative anxiety management, followed by retrospective analysis of prospectively collected implementation data. Our study objectives were to: (1) evaluate the effectiveness of virtual reality intervention on pediatric preoperative distress, fear, and physiological markers; (2) identify predictors of clinically significant anxiety reduction; (3) assess stakeholder experience including patient satisfaction, parent anxiety, and healthcare provider acceptance; and (4) examine factors associated with successful implementation defined as combined clinical response and stakeholder satisfaction.

## MATERIALS AND METHODS

### Ethics Statement

This study was approved by the Virginia Commonwealth University Institutional Review Board (Protocol #HM20024981, NCT07280910) and classified as non-human subjects research involving retrospective analysis of de-identified data collected during a clinical quality improvement initiative. The quality improvement project was conducted under standard hospital operational protocols at VCU Children’s Hospital. During the quality improvement implementation phase, written informed consent was obtained from parents or legal guardians and age-appropriate assent from pediatric participants prior to virtual reality intervention, following detailed explanation of procedures, potential risks, benefits, and participant rights. All consent documentation was written at an eighth grade reading level to ensure comprehension across diverse literacy levels. The study was conducted in accordance with the Declaration of Helsinki and International Conference on Harmonisation Good Clinical Practice guidelines.

### Study Design and Setting

We conducted a retrospective analysis of prospectively collected data generated during a nine-month quality improvement initiative (October 2023 through June 2024) implementing virtual reality interventions for pediatric preoperative anxiety management. The initiative took place at VCU Children’s Hospital in Richmond, Virginia, specifically within the pediatric preoperative holding area, a 16-bed unit managing approximately 1,000 surgical cases annually. The study employed a single-arm, pre-post design to examine within-subject changes in psychological and physiological outcomes associated with virtual reality intervention, while characterizing stakeholder experience and testing predictors of clinically meaningful improvement under routine clinical conditions.

### Quality Improvement Framework

The implementation followed Plan-Do-Study-Act methodology across five complete cycles, each lasting 4-6 weeks. This iterative approach enabled continuous refinement of virtual reality intervention protocols based on real-time data collection and stakeholder feedback. Each cycle followed structured processes of planning interventions based on previous findings, implementing changes in clinical practice, studying outcomes through systematic data collection and analysis, and acting on results to standardize successful modifications. The initiative aimed to: (1) implement virtual reality technology as non-pharmacological intervention for pediatric preoperative anxiety; (2) optimize clinical workflow integration; (3) maximize satisfaction among patients, parents, and healthcare providers; and (4) continuously refine protocols based on stakeholder feedback and outcomes.

Cycle 1 (October-November 2023) established feasibility and safety through standardized 3-5 minute intervention sessions with systematic documentation of adverse events, setup time, technical failures, and age-stratified usability assessment. Cycle 2 (January-February 2024) introduced tablet-based parental viewing capability while establishing baseline effectiveness metrics with standardized five-minute sessions. Cycle 3 (March-April 2024) implemented flexible session duration (5-20 minutes) based on patient preference and expanded clinical applications beyond procedural support to general preoperative anxiety management. Cycle 4 (May 2024) validated the refined protocol with minimum 10-minute session duration and indication-specific implementation. Cycle 5 (June 2024) completed data collection while assessing operational sustainability and staff competency.

### Study Population and Eligibility

#### Pediatric participants

Eligible patients were children aged 6-15 years scheduled for elective surgical procedures, capable of understanding instructions and providing self-report on validated anxiety scales, able to wear a virtual reality headset safely, and accompanied by a parent or legal guardian able to provide informed consent. Exclusion criteria included history of seizures or severe motion sickness, sensory or cognitive impairment preventing safe virtual reality use, active infection or open wounds on head or face, positive influenza-like illness screen on day of surgery, and any additional contraindication identified at clinician discretion.

#### Parent or caregiver participants

Parents or legal guardians present during the preoperative period and willing to complete anxiety and satisfaction measures were eligible for participation.

#### Healthcare provider participants

Preoperative nurses involved in virtual reality setup and supervision and willing to complete satisfaction and feasibility ratings were included.

Participants were recruited using convenience sampling during routine preoperative visits. The nursing team identified eligible patients from daily surgical schedules and notified the quality improvement team of potential participants. Initial recruitment occurred three days weekly (Tuesday, Wednesday, Friday), expanding to five days weekly with extended hours by Cycle 3 based on patient volume analysis.

### Virtual Reality Intervention

#### Technology selection

The Meta Quest 2 virtual reality headset was selected following systematic evaluation of available technologies against criteria including pediatric safety profile, ease of disinfection between patients, age-appropriate content availability, cost-effectiveness, and technical support requirements. The device offered wireless functionality important for the clinical environment and met all safety and practical requirements.

#### Content development

Custom virtual reality environments were developed over 10 months (November 2022 through August 2023) using Unity 3D software, incorporating evidence-based design principles for pediatric anxiety reduction. The environment featured an interactive children’s room containing familiar toys and objects responsive to gaze and controller input, virtual television displaying age-appropriate content, warm color palette (predominantly blues and greens) associated with reduced anxiety, spatial audio with background music at 60-70 beats per minute, and consistent lighting to minimize visual fatigue. A secondary beach scene featured gentle ocean waves, natural elements including palm trees and cloud movements, and calming environmental sounds.

To minimize motion-related discomfort, the environment employed fixed viewpoint design where users remained in stable virtual locations with teleportation-based navigation allowing point-to-point movement between specific locations without experiencing visual acceleration. This approach minimized vestibular-visual conflict, a primary factor in motion-related discomfort particularly relevant to pediatric populations. Effectiveness of this design was confirmed through pilot testing with 15 pediatric patients, with zero reports of motion discomfort or nausea.

#### Intervention protocol

Following quality improvement refinement cycles, the final protocol consisted of: (1) eligibility screening by Child Life Specialist; (2) informed consent and assent procedures; (3) baseline assessment including demographic information, physiological measures from bedside monitors, and self-reported distress and fear using validated scales; (4) virtual reality headset fitting and orientation; (5) patient-directed virtual reality session with minimum 10-minute duration and maximum 20-minute duration based on patient preference and clinical workflow; (6) concurrent parental viewing via tablet-based mirroring system; (7) immediate post-intervention assessment of distress, fear, and physiological parameters; and (8) satisfaction feedback from patients, parents, and nursing staff.

### Outcome Measures

#### Primary outcomes

Procedural distress was measured using a 0–10 Numerical Rating Scale (NRS), where 0 indicated “no distress” and 10 indicated “extreme distress.” Although originally validated for pediatric pain, the NRS has also demonstrated validity for assessing distress and emotional responses during medical procedures. Fear was assessed using the Children’s Fear Scale (CFS), a 5-face pictorial measure scored 0–4, validated for children aged 7–12 years in procedural contexts. Both measures were administered at baseline (T1) and immediately post-intervention (T2) using standardized prompts.

#### Secondary outcomes

Physiological parameters included heart rate (beats/min) and systolic blood pressure (mmHg), recorded five minutes pre-intervention and immediately post-intervention using calibrated bedside monitors. Parent anxiety was measured using the 6-item State-Trait Anxiety Inventory short form (STAI-6), which provides a brief, validated measure of situational anxiety. Parents completed the STAI-6 at baseline and post-intervention.

#### Satisfaction and feasibility

Satisfaction was assessed using 5-point Likert scales (1=“extremely dissatisfied” to 5=“extremely satisfied”). Children and parents independently rated intervention satisfaction and overall procedural experience. Nurses completed parallel scales evaluating feasibility, satisfaction, and perceived effectiveness in anxiety management.

### Data Collection Procedures

Patient recruitment was facilitated by the unit’s Child Life Specialist who conducted initial eligibility screening and provided information to parents and patients. Eligible patients and accompanying parents were referred to the research team for enrollment. Research personnel explained the study, obtained written informed consent, and proceeded with baseline data collection.

Pre-intervention assessment included demographic information (age, gender), baseline physiological measures (heart rate, systolic blood pressure) recorded from bedside monitors, and participant self-reported baseline distress and fear assessed using validated scales. For parents consenting to participate, baseline state anxiety was measured using the State-Trait Anxiety Inventory-6.

During virtual reality intervention, process metrics were documented including session start and end times to calculate total duration, primary clinical indication for use (procedural support versus general anxiety), and any observed adverse effects or technical issues. Upon virtual reality completion, immediate post-intervention assessment was conducted involving readministration of distress and fear scales, post-intervention heart rate and blood pressure recorded from monitors, participant feedback via satisfaction scales, and indication of willingness to use virtual reality in future procedures. Participating parents simultaneously completed post-intervention anxiety measurements and provided feedback regarding their perceptions of intervention effectiveness. Nursing staff completed implementation feasibility and satisfaction assessments.

### Data Management and Quality Assurance

All data were de-identified at the point of collection using participant codes and stored in a secure, password-protected database. Data entry was checked against original forms to ensure accuracy, and the dataset was reviewed for out-of-range values and logical inconsistencies. Research staff verified completeness of data forms at the time of collection. Regular audits and monitoring ensured >95% data completeness and integrity across all study variables.

### Statistical Analysis

Analyses were conducted using Python (v3.12). Descriptive statistics summarized baseline demographic and clinical variables. Normally distributed variables (e.g., age, heart rate, blood pressure) were summarized as means (SD) and compared using paired t-tests. Non-normally distributed variables (distress and fear) were summarized as medians (IQR) and compared using Wilcoxon signed-rank tests. Clinically significant improvement was defined a priori as ≥30% reduction from baseline scores, consistent with pediatric anxiety and pain research. Effect sizes were estimated using change scores and 95% CIs.

Logistic regression models examined predictors of clinically significant distress or fear reduction (≥50%). Predictor variables included age, baseline scores, and VR session duration. Odds ratios with 95% CIs were reported. Parent anxiety (STAI-6) was compared pre–post using Wilcoxon tests. Parent, patient, and nurse satisfaction were summarized descriptively; associations between parent and child acceptance were tested with chi-square tests.

Implementation success was defined as a composite outcome requiring ≥30% reduction in distress or fear plus high parental satisfaction (≥4/5). Predictors were examined using logistic regression. Model fit was evaluated with pseudo-R² and likelihood ratio tests. Qualitative analysis of parent feedback employed thematic analysis. Sensitivity analyses tested robustness using a ≥50% reduction threshold and by excluding outliers.

### Sample Size Determination

Sample size was determined a priori based on expected reductions in distress and fear. Prior VR studies in pediatric populations have reported moderate effects (SMD ≈0.5). Assuming a small-to-moderate effect (Cohen’s dz=0.35), two-tailed α=0.05, and 80% power, we estimated that 67 pairs were required for parametric testing, adjusted to 71 for non-parametric tests. Allowing for ∼10% attrition, the target enrollment was 80 participants. For secondary logistic regression analyses, we applied the rule of ≥10 events per predictor variable. With approximately 40 expected responders, this sample was adequate to support up to four predictors.

### Handling of Missing Data

Missing data were minimal (<5% across all variables). Analyses were performed using complete case analysis, retaining only participants with data available for all variables in a given test. Change scores and binary clinical response variables were computed according to the pre-specified analysis plan.

## RESULTS

### Participant Characteristics

A total of 80 pediatric patients undergoing preoperative procedures were enrolled between October 2023 and June 2024. The mean age of pediatric patients was 10.4±3.0 years (range 6-15 years), with higher proportion of females (68.8%, 55/80) than males (31.2%, 25/80). The virtual reality intervention duration averaged 9.3±4.2 minutes with median of 9.0 minutes (interquartile range 6.0-11.2 minutes, range 5-20 minutes).

Among 65 parent or caregiver participants, most were mothers (81.5%, 53/65) with remainder being fathers (18.5%, 12/65). The majority of parents (58.5%, 38/65) were between 31-40 years of age. Educational background varied, with largest group (38.5%, 25/65) having university or professional degrees. Almost two-thirds (64.6%, 42/65) reported previous emergency department visits with their child. Among 29 participating nurses, all completed satisfaction and feasibility assessments. Participant demographic characteristics are summarized in Table 1.

**Table 1.** Demographic Characteristics of Study Participants.

| Characteristics | N (%) or Mean (SD) |
| --- | --- |
| **Study Participants (N=80)** |  |
| Age, years (mean ± SD) | 10.4 ± 3.0 |
| Age range (years) | 6–15 |
| **Gender, n (%)** |  |
| Male | 25 (31.2%) |
| Female | 55 (68.8%) |
| **VR Duration (min)** |  |
| Mean (SD) | 9.3 ± 4.2 |
| Median (IQR) | 9.0 (6.0-11.2) |
| Range | 5-20 |
| **Parent/Caregiver Characteristics (N=65)** |  |
| **Relationship to Child, n(%)** |  |
| Mother | 53(81.5%) |
| Father | 12(18.5%) |
| **Parent Age, n (%)** |  |
| 20-30 years old | 11 (16.9%) |
| 31-40 years old | 38 (58.5%) |
| 41-50 years old | 10 (15.4%) |
| 51-60 years old | 6 (9.2%) |
| **Parent Education, n (%)** |  |
| University/ Professional Degree | 25 (38.5%) |
| Diploma/Certificate | 18 (27.7%) |
| High school | 10 (15.4%) |
| Some Post Secondary University | 10 (15.4%) |
| Elementary school | 2 (3.1%) |
| **Prior ED visit** |  |
| Yes | 42 (64.6%) |
| No | 23 (35.4%) |
| *Abbreviations: ED = Emergency Department; IQR = Interquartile Range; SD = Standard Deviation; VR = Virtual Reality* |  |

### Effect of Virtual Reality on Distress and Fear

Distress scores decreased significantly from median 3.0[2.0–5.0] pre-intervention to median 1.0 [0.0–2.2] post-intervention, with median change of -2.0 (Wilcoxon signed-rank test W=138.5, p<0.0001). Among children with baseline scores greater than zero, 79.1% (53/67; 95% confidence interval 67.4-88.1) achieved the predefined clinically significant threshold of ≥30% reduction in distress. Using an absolute threshold of ≥2-point reduction to include baseline-zero cases yielded 68.8% response rate (55/80; 95% confidence interval 57.4-78.7).

Fear scores showed significant improvement, decreasing from median 1.0[0.0–2.0] to median 0.0 [0.0–1.0] with median change of -1.0 (Wilcoxon signed-rank test W=186.0, p<0.001). Over half of participants (52.5%; 95% confidence interval [41.0-63.8]) achieved ≥30% reduction in fear. Pre-post comparisons of psychological outcome measures are presented in Table 2, and clinical significance achievement rates are shown in Table 3.

**Table 2.** Pre-Post Comparison of Psychological Outcome Measures.

| **Table 2. Pre-Post Comparison of Psychological Outcome Measures** |  |  |  |  |
| --- | --- | --- | --- | --- |
| **Outcome Measure** | **Pre-VR Median [IQR]** | **Post-VR Median [IQR]** | **Median Change (95% CI)** | **P-value\*** |
| **Distress Score (0-10)** | 3.0[2.0–5.0] | 1.0 [0.0–2.2] | - 2.0 (2.00, 1.00) | <0.001 |
| **Fear Score (0-4)** | 1.0[0.0–2.0] | 0.0 [0.0–1.0] | -1.0 (0.00, 1.00) | <0.001 |
| *\*Wilcoxon signed-rank test for non-parametric paired comparisons*<br>Abbreviations: CI, confidence interval; IQR, interquartile range; VR, virtual reality |  |  |  |  |

**Table 3.** Clinical Significance Achievement by Outcome.

| **Table 3. Clinical Significance Achievement by Outcome** |  |  |
| --- | --- | --- |
| **Outcome** | **Patients Achieving $\geq 30\%$ Reduction n/n (%)** | **95% CI** |
| Distress | 53/80 (66) | 55 - 75 |
| Fear | 42/80 (52) | 41 - 63 |
| *Abbreviations: CI = confidence interval* |  |  |

### Physiological Outcomes

Analysis of physiological parameters revealed significant reductions following virtual reality intervention. Mean heart rate decreased from 86.3±14.7 beats per minute pre-intervention to 80.5±12.7 beats per minute post-intervention, resulting in mean reduction of 5.8±13.0 beats per minute. This decrease was statistically significant (paired t-test t=3.95, p<0.001). Similarly, systolic blood pressure demonstrated significant reduction from baseline 113.9±10.6 mmHg to post-intervention 109.8±10.9 mmHg, with mean decrease of 4.1±8.5 mmHg (paired t-test t=4.31, p<0.0001). Changes in physiological markers are presented in Table 4.

**Table 4.**
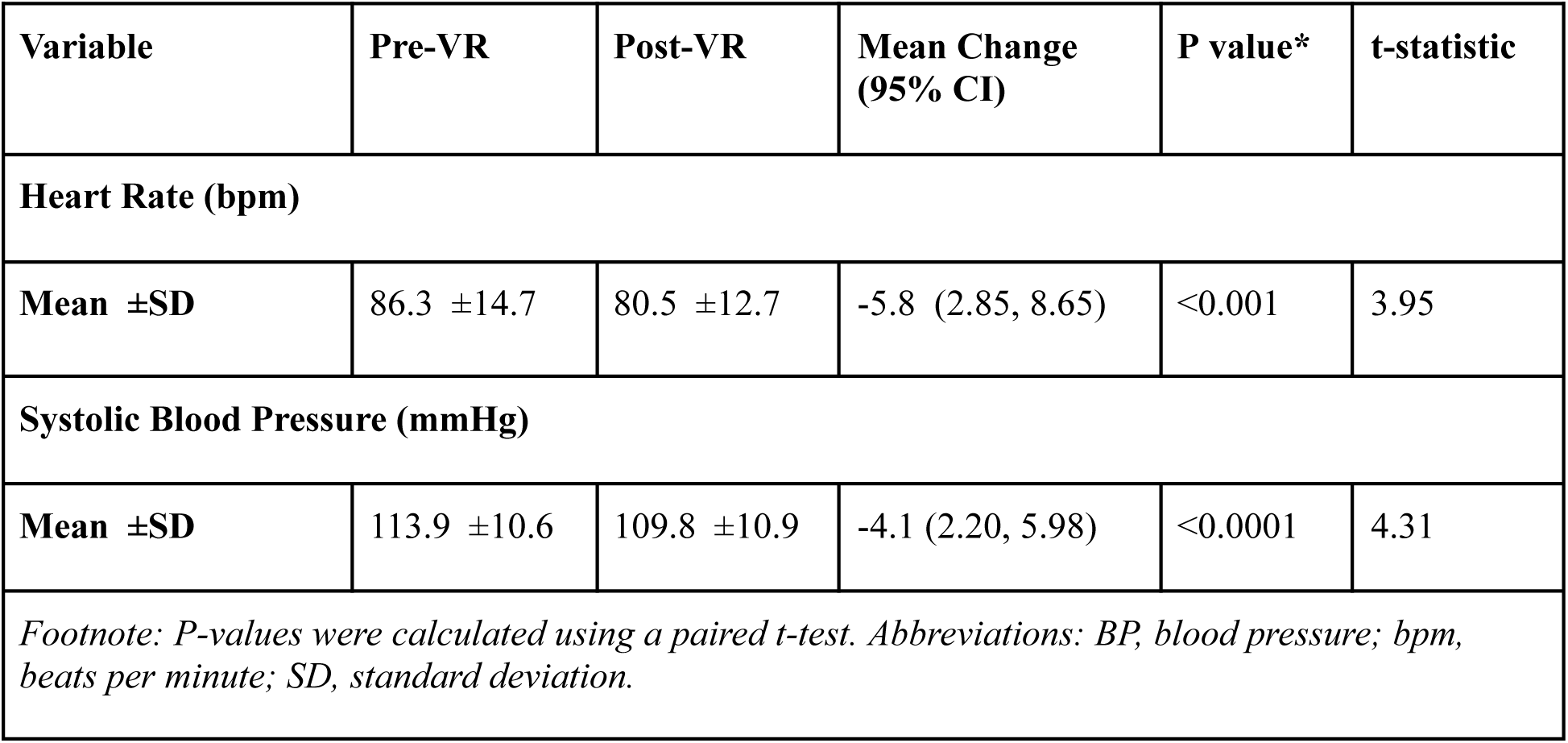
Changes in Physiological Markers Following VR Intervention.

| **Table 4. Changes in Physiological Markers Following VR Intervention** |  |  |  |  |  |
| --- | --- | --- | --- | --- | --- |
| **Variable** | **Pre-VR** | **Post-VR** | **Mean Change**<br>**(95% CI)** | **P value\*** | **t-statistic** |
| **Heart Rate (bpm)** |  |  |  |  |  |
| **Mean ±SD** | 86.3 ±14.7 | 80.5 ±12.7 | -5.8 (2.85, 8.65) | <0.001 | 3.95 |
| **Systolic Blood Pressure (mmHg)** |  |  |  |  |  |
| **Mean ±SD** | 113.9 ±10.6 | 109.8 ±10.9 | -4.1 (2.20, 5.98) | <0.0001 | 4.31 |
| *Footnote: P-values were calculated using a paired t-test. Abbreviations: BP, blood pressure; bpm, beats per minute; SD, standard deviation.* |  |  |  |  |  |

### Predictors of Clinical Response

Two separate logistic regression models examined predictors of clinically significant reduction (≥50% improvement) in fear and distress. The model for fear reduction was statistically significant overall (log-likelihood ratio p-value=0.038, pseudo R²=0.038). Baseline fear score emerged as the only statistically significant predictor of substantial fear reduction (odds ratio 1.80, 95% confidence interval 1.11-2.92, p=0.017), indicating that for each one-point increase in baseline fear score, the odds of achieving clinically significant reduction increased by 80%. Neither age (odds ratio 0.89, 95% confidence interval 0.75-1.05, p=0.162) nor virtual reality duration (odds ratio 0.99, 95% confidence interval 0.88-1.11, p=0.832) demonstrated significant associations with likelihood of fear reduction.

The model for distress reduction was statistically significant (log-likelihood ratio p-value=0.040, pseudo R²=0.076). Baseline distress score was identified as significant predictor (odds ratio 1.33, 95% confidence interval 1.06-1.66, p=0.014), indicating that each one-point increase in baseline distress was associated with 33% increase in odds of achieving clinically significant reduction. As with the fear model, neither age (odds ratio 0.92, 95% confidence interval 0.78-1.09, p=0.362) nor virtual reality duration (odds ratio 0.99, 95% confidence interval 0.88-1.11, p=0.829) significantly predicted distress reduction. Predictor analyses are presented in Table 5.

**Table 5.** Logistic Regression Analysis: Predictors of Clinical Response to VR Intervention.

| **A. Predictors of Fear Reduction ($\geq 50\%$ improvement)** |  |  |  |  |  |
| --- | --- | --- | --- | --- | --- |
| **Variable** | **$\beta$ Coefficient** | **SE** | **OR (95% CI)** | **P-value** | **Model Statistics** |
| Intercept | -0.16 | 1.17 | - | 0.890 |  |
| Age (years) | -0.12 | 0.09 | 0.89 (0.75, 1.05) | 0.162 |  |
| Baseline Fear (points) | 0.59 | 0.25 | 1.80 (1.11, 2.92) | *0.017 | Pseudo $R^2 = 0.038$ |
| VR Duration (min) | -0.01 | 0.06 | 0.99 (0.88, 1.11) | 0.832 | LLR $p$-value:<br>0.038 |
| **B. Predictors of Distress Reduction ($\geq 50\%$ improvement)** |  |  |  |  |  |
| **Variable** | **$\beta$ Coefficient** | **SE** | **OR (95% CI)** | **P-value** | **Model Statistics** |
| Intercept | 0.32 | 1.22 | - | 0.789 |  |
| Age (years) | -0.076 | 0.08 | 0.92 (0.78, 1.09) | 0.362 |  |
| Baseline Distress (points) | 0.273 | 0.11 | 1.33 (1.06, 1.66) | *0.014 | Pseudo R<sup>2</sup> = 0.076 |
| VR Duration (min) | -0.012 | 0.06 | 0.99 (0.88, 1.11) | 0.829 | LLR p-value: 0.040 |
| *Abbreviations: CI = confidence interval; LLR = log-likelihood ratio; OR = odds ratio; SE = standard error. Model fit: N=80; Responders=46; Non-responders=34; Predictors=3; EPV(min)=11.3; Pseudo R<sup>2</sup>=0.076; LLR test p=0.040; Converged=Yes.*<br>*Notes: Outcome = indicator of ≥50% reduction in distress from baseline to post-VR. Odds ratios from logistic regression with HC3 robust standard errors. OR>1 indicates higher odds of meeting the responder threshold per 1-unit increase in the predictor.* |  |  |  |  |  |

### Stakeholder Experience

#### Parent and caregiver outcomes

Analysis of parent and caregiver outcomes (n=65) revealed consistently positive responses to virtual reality intervention. Parent satisfaction with procedure was high, with mean score of 4.55±0.69 on 5-point scale (95% confidence interval 4.38-4.72), indicating strong approval of overall procedural experience. Satisfaction with anxiety management was rated at 4.18±0.90 (95% confidence interval 3.96-4.40). Notably, 95.4% of parents and caregivers (62/65) indicated they would recommend virtual reality intervention for other children undergoing similar procedures (95% confidence interval 87.1-99.0%).

#### Pediatric patient outcomes

Pediatric patients reported high levels of satisfaction with virtual reality intervention. Mean overall happiness score was 4.34±0.83 on 5-point scale (95% confidence interval 4.16-4.52). Satisfaction ratings were predominantly positive, with 86.2% reporting high (score=4) or very high (score=5) satisfaction. Only 2.4% of participants reported low satisfaction (scores 1-2). Large majority of children (91.2%, 73/80) reported willingness to try virtual reality again in future procedures (95% confidence interval 82.8-96.4%), while 88.8% (71/80) specifically indicated that virtual reality helped with their anxiety (95% confidence interval 79.7-94.7%). Adverse effects were minimal, occurring in only 3.8% (3/80, 95% confidence interval 0.8-10.6%) of participants.

#### Healthcare provider outcomes

Nursing staff (n=29) reported positive responses to integrating virtual reality into pediatric procedural care. The majority of nurses (82.8%) rated the virtual reality system as “extremely easy” to manage during clinical procedures (mean implementation ease score 4.72±0.53 on 5-point scale). Satisfaction among nursing staff was consistently high, with 75.9% (22/29) reporting being “very satisfied” and 17.2% (5/29) “satisfied” with intervention. No nurses reported dissatisfaction with virtual reality implementation. Notably, all participating nurses (100%, 29/29) indicated they would recommend virtual reality for pediatric procedures. Only 6.9% (2/29) of nurses reported any workflow disruption associated with intervention, while 93.1% (27/29) reported that virtual reality enhanced patient care. Stakeholder satisfaction and acceptance outcomes are presented in Table 6.

**Table 6.** Stakeholder Satisfaction and Acceptance Outcomes.

| **Table 6. Stakeholder Satisfaction and Acceptance Outcomes** |  |  |
| --- | --- | --- |
| **A. Parent/Caregiver Experience (n=65)** |  |  |
| **Outcome Measure** | **n (%) or Mean ±SD** | **95% CI** |
| Procedure Satisfaction (1-5) | $4.55 \pm 0.69$ | (4.38, 4.72) |
| Anxiety Management Satisfaction (1-5) | $4.18 \pm 0.90$ | (3.96, 4.40) |
| Would Recommend VR (n, %) | 62/65 (95%) | (87%, 99%) |

| **B. Pediatric Patient Experience (n=80)** |  |  |
| --- | --- | --- |
| **Outcome Measure** | **n (%) or Mean ±SD** | **95% CI** |
| Overall Happiness Score (1-5) | $4.34 \pm 0.83$ | (4.16, 4.52) |
| Would Try VR Again(n, %) | 73/80 (91%) | (82%, 96%) |
| VR Helped with Anxiety | 71/80 (88 %) | (79%, 94%) |
| Experienced Side Effect | 3/80 (3%) | (0.8%, 10%) |

| **C. Nursing Staff Experience (n=29)** |  |
| --- | --- |
| **Outcome Measure** | **n (%) or Mean (SD)** |
| Ease of Implementation (1-5) | 4.72 ±0.53 |
| Overall Satisfaction (1-5) | 4.69 ±0.60 |
| Would Recommend VR | 29/29 (100%) |
| Workflow Disruption | 2/29 (6%) |
| Enhanced Patient Care | 27/29 (93%.) |
| *Statistically significant reduction (p < 0.01) |  |

### Parent Anxiety Outcomes

Analysis of parental anxiety revealed statistically significant reduction following virtual reality intervention (p<0.0015). Median State-Trait Anxiety Inventory-6 score pre-intervention was 14.0 (interquartile range 11.0-17.0) and remained 14.0 post-intervention but with narrower distribution (interquartile range 11.0-15.0). Median change in anxiety score was -1.0 (interquartile range -4.00 to 0.00) with 95% confidence interval of -2.00 to 0.00.

Distribution of change scores indicated that 53.8% (35/65) of parents experienced improved anxiety levels following their child’s virtual reality intervention, while 23.1% (15/65) showed no change and 23.1% (15/65) reported increased anxiety. Parent anxiety outcomes are presented in Table 7.

**Table 7.** Parent/Caregiver anxiety (n=65)

| **Table7. Parent/Caregiver anxiety (n=65)** |  |  |  |
| --- | --- | --- | --- |
| **Outcome Measure** | **Median [IQR]** | **95% CI** | **P-value** |
| Pre-VR Anxiety Score | 14 [11–17] | [13, 15] | <0.0015 |
| Post-VR Anxiety Score | 14 [11 –15] | [12, 15] |  |
| Median Change | -1 [-4 - 0] | [-2, 0] |  |

### Parent-Child Concordance in Virtual Reality Acceptance

Chi-square test of independence examined association between parent’s willingness to recommend virtual reality and child’s willingness to use it again. Results indicated no statistically significant association between the two (χ²(1)=0.11, p=0.74). In 57 of 65 dyads (87.7%), parent and child responses were concordant. Most notably, 56 dyads (86.2%) demonstrated mutual positive acceptance of intervention. Discordant responses were relatively uncommon, with 6 cases (9.2%) where children were willing to use virtual reality again but parents would not recommend it, and 2 cases (3.1%) where parents would recommend virtual reality despite child’s unwillingness to use it again. Contingency analysis is presented in Table 8.

**Table 8.** Contingency Table of Parent and Child VR Acceptance.

| **Table 8. Contingency Table of Parent and Child VR Acceptance** |  |  |
| --- | --- | --- |
|  | **Child Willing to Use VR Again** |  |
| **Parent Recommends VR** | No | Yes |
| No | 1 | 2 |
| Yes | 6 | 56 |

### Qualitative Findings: Parent Perspectives

Thematic analysis of open-ended feedback from 63 parents yielded six distinct themes regarding perceptions of virtual reality intervention. Engagement and Distraction emerged as most prevalent theme, present in 52.4% (33/63) of parent responses. This theme encompassed comments about virtual reality’s capacity to capture children’s attention and divert it from clinical procedures. As one parent noted, “He was distracted and didn’t notice when the needle went in,” while another commented, “It takes away the attention of the child.”

Emotional Impact was identified in 34.9% (22/63) of responses, making it second most frequent theme. Parents described virtual reality intervention’s effect on reducing anxiety and promoting calm for both children and parents. One parent of child with special needs observed, “He has autism and he seems much more relaxed while wearing the VR. He’s not thinking about the procedure while waiting.” Another simply stated, “Very cool to watch, helpful to know my child was not in pain.”

Clinical Utility appeared in 31.7% (20/63) of parent comments, focusing on practical benefits of virtual reality in healthcare setting. Parents recognized broader applications beyond their individual experience, with comments such as “will help a lot of kids and their parents” and “It helps remove the thoughts of over thinking and worry.”

Stakeholder Experience (19.0%, 12/63) captured distinct perspectives of different individuals involved in virtual reality implementation. Some parents contrasted their own experience with their child’s: “If my child chose it, sure. To me it’s a bit overwhelming to have so much going on.”

Challenges and Concerns were mentioned in only 4.8% (3/63) of responses, primarily from parents reporting neutral satisfaction. These comments addressed implementation difficulties or perceived limitations: “I think it was difficult to get a vein because the skin was thick, and it hurt.”

Technology Experience was least frequent theme (3.2%, 2/63), with limited comments specifically addressing virtual reality technology itself rather than its effects: “This headset was a great idea.”

Analysis of theme co-occurrence revealed that Clinical Utility and Engagement and Distraction frequently appeared together (9 responses), suggesting parents connected virtual reality’s ability to distract with its practical clinical value. Similarly, Engagement and Distraction frequently co-occurred with Stakeholder Experience (9 responses), indicating parents often framed observations of virtual reality intervention in terms of their child’s engagement and their own observations. Thematic analysis summary is presented in Table 9.

**Table 9.** Thematic Analysis of Parent Perspectives on VR Intervention (N=63)

| Theme | Description | Prevalence | Representative Quotation |
| --- | --- | --- | --- |
| Engagement & Distraction | VR's capacity to capture attention, provide distraction from medical procedures, and create an engaging experience | 33 (52%) | "He was distracted and didn't notice when the needle went in" |
| Emotional Impact | Effect on emotional states, including reduction of anxiety, fear, and promotion of calm | 22 (35%) | "He has autism, and he seems much more relaxed while wearing the VR. He's not thinking about the procedure while waiting" |
| Clinical Utility | Practical benefits and applications in the clinical setting | 20 (32%) | "Will help a lot of kids and their parents" |
| Stakeholder Experience | Distinct experiences and perspectives of different stakeholders | 12 (19%) | "If my child chose it, sure. To me it's a bit overwhelming to have so much going on" |
| Technology Experience | Perceptions of the VR technology itself | 2 (3%) | "This headset was a great idea" |

### Predictors of Implementation Success

For composite outcome of Implementation Success (defined as clinical improvement of ≥30% plus positive stakeholder feedback), logistic regression model identified key predictors. The overall model was statistically significant (pseudo R²=0.118, p=0.017). Analysis revealed that baseline fear score was sole significant predictor. For each one-point increase in child’s initial fear score, odds of achieving successful implementation increased by 130% (odds ratio 2.30, 95% confidence interval 1.09-4.84, p=0.029). Neither baseline distress score (odds ratio 1.07, 95% confidence interval 0.83-1.37, p=0.609) nor age (odds ratio 0.92, 95% confidence interval 0.76-1.12, p=0.415) were significant predictors in this model. Implementation success predictors are presented in Table 10.

**Table 10:** Predictors of Implementation Success (Logistic Regression)

| **Table 1: Predictors of Implementation Success (Logistic Regression)** |  |  |  |  |  |
| --- | --- | --- | --- | --- | --- |
| **Variable** | **$\beta$ Coefficient** | **SE** | **OR (95% CI)** | ***P* value** | **Model Statistics** |
| Age | -0.079 | 0.097 | 0.92(0.76, 1.12) | 0.415 |  |
| Baseline Distress Score | 0.066 | 0.129 | 1.07(0.83, 1.37) | 0.609 | Pseudo R-squared: 0.118 |
| Baseline Fear Score | 0.831 | 0.381 | 2.30(1.09, 4.84) | 0.029 | LLR p-value: 0.017 |
| *Abbreviations: CI = confidence interval; LLR = log-likelihood ratio; OR = odds ratio; SE = standard error* |  |  |  |  |  |

### Sensitivity Analyses

Two sensitivity analyses assessed robustness of primary findings. First, analysis defining clinical significance was repeated using stricter threshold of ≥50% reduction from baseline. Using this more rigorous criterion, 46 of 80 patients (57.5%, 95% confidence interval 46.6-67.7%) achieved clinically significant reduction in distress, and 36 of 80 patients (45.0%, 95% confidence interval 34.6-55.9%) achieved this for fear.

Second, robustness check was performed by re-running primary Wilcoxon signed-rank tests after excluding 19 participants who were potential outliers at age extremes or with very short or long virtual reality durations. Even with this smaller, more homogenous sample (n=61), significant reductions in both distress (median change=-2.0, p<0.0001) and fear (median change=-1.0, p<0.0001) remained. These analyses demonstrated that study’s primary findings were robust, not dependent on specific definition of clinical significance, and not driven by outlier participants.

## DISCUSSION

This study demonstrated that immersive virtual reality intervention significantly reduced preoperative anxiety in pediatric patients while achieving high acceptance across children, parents, and healthcare providers. The findings suggest comprehensive evidence supporting virtual reality integration into routine pediatric perioperative care protocols, addressing both clinical effectiveness and implementation feasibility under real-world conditions.

### Clinical Effectiveness and Underlying Mechanisms

The primary outcomes strongly supported hypotheses that virtual reality would reduce children’s distress and fear in the preoperative holding period. Median self-reported distress scores decreased from 3 to 1 on the 0-10 scale, and fear scores dropped from 1 to 0 on the 0-4 pictorial scale, both representing highly significant improvements. In practical terms, two-thirds of children achieved clinically meaningful reduction (≥30%) in subjective distress, and over half achieved this level of reduction in fear aligning with established clinical significance thresholds and prior pediatric VR meta-analyses.

The significant physiological improvements observed provide objective validation of subjective anxiety reductions reported by pediatric participants, demonstrating that virtual reality intervention produces measurable autonomic nervous system changes rather than simple reporting bias. The 5.8 beats per minute reduction in heart rate and 4.1 mmHg decrease in systolic blood pressure reflect decreased sympathetic nervous system activation and enhanced parasympathetic tone, indicating genuine stress reduction rather than merely distraction effects. Recent pediatric virtual reality studies have documented similar physiological benefits, with investigations reporting significant reductions in electrodermal activity (68% decrease) and blood volume pulse amplitude (53% decrease) during virtual reality exposure in dental procedures, as well as comparable heart rate reductions in virtual reality versus control groups during medical procedures (10,11). These autonomic improvements are particularly significant in pediatric populations where anxiety-induced cardiovascular activation can complicate anesthesia induction, increase procedural risk, and prolong recovery.

The physiological validation contributes to the growing evidence for virtual reality as non-pharmacological intervention producing measurable, objective benefits beyond subjective comfort improvements, supporting its integration into routine pediatric perioperative care protocols(12,13).

### Predictors of Treatment Response

The study identified factors associated with greater benefit from virtual reality intervention, with baseline anxiety levels emerging as primary predictors of clinical response. Children with higher initial distress and fear scores experienced the largest improvements. For each one-point increase in baseline fear (0-4 scale), odds of marked fear reduction doubled (odds ratio 1.8), and similarly each additional point of baseline distress (0-10 scale) raised odds of major distress relief by 33%. These findings suggest that virtual reality was especially effective for most anxious children, who arguably have most to gain from distraction intervention. This pattern aligns with established treatment response literature indicating that individuals with higher baseline symptom severity demonstrate greater capacity for improvement, thereby enhancing their probability of achieving clinically significant reductions(14–16).

Age did not emerge as significant moderator of virtual reality effectiveness across the 6-15 year range, contrasting with mixed findings in existing literature (7,13,17). While some studies suggest younger children (4-8 years) may show greater anxiety reductions during needle procedures, other research including meta-analytic evidence found no significant age moderation of virtual reality effectiveness for pediatric anxiety during needle procedures within 4-12 year age range. Our results support the conclusion that virtual reality’s immersive features can effectively reduce anxiety across children aged 6 to 15 years. However, because we included wide age range, it is possible that age-related differences in how children respond to intervention were not fully captured.

Regarding session duration (5-20 minutes), lack of linear dose-response relationship once baseline anxiety was controlled suggests that beyond minimum threshold engagement time, simply using virtual reality was broadly effective. Professional guidelines recommend 10-15 minute sessions for school-age children to optimize benefits while minimizing potential adverse effects. Notably, in earlier quality improvement cycles longer sessions (≥10 minutes) tended to yield somewhat larger anxiety reductions on average, suggesting that minimum threshold of engagement around 10 minutes might be optimal. In practice, allowing children to use virtual reality for as long as they desire within workflow constraints appears reasonable.

### Stakeholder Acceptance and System Integration

The comprehensive stakeholder outcomes demonstrate multi-level implementation success for virtual reality intervention, reflecting alignment of positive experiences across all key healthcare participants. Consistently high satisfaction ratings across parents and caregivers (mean 4.55±0.69), pediatric patients (mean 4.34±0.83), and nursing staff (mean 4.72±0.53) align with recent implementation studies showing that successful virtual reality integration requires stakeholder alignment and acceptance(18,19). High level of parental willingness to recommend virtual reality to other families (95.4%) and children’s expressed desire to use virtual reality again (91.2%) mirrors findings from other pediatric virtual reality implementation studies, where high satisfaction rates and positive user experiences drive sustained adoption and clinical integration.

Healthcare provider acceptance, with 100% of nurses recommending virtual reality and 93.1% reporting enhanced patient care, reflects critical implementation facilitators identified in multi-stakeholder research, including ease of use, minimal workflow disruption, and observed patient benefits. The significant reduction in parental anxiety (p<0.001) with 53.8% of parents experiencing improved anxiety levels following their child’s virtual reality intervention confirms recent studies demonstrating that family-centered virtual reality interventions can effectively address caregiver distress in pediatric healthcare settings. Our finding that parents had significantly lower state anxiety after virtual reality session represents important but underexplored aspect of pediatric virtual reality interventions, suggesting that virtual reality not only helps child directly but also indirectly calms caregiver by alleviating distress of watching one’s child in anxiety.

The high degree of parent-child concordance in virtual reality acceptance (87.7% agreement) with predominantly mutual positive responses (86.2% of dyads) suggests successful alignment between patient and family preferences, critical factor in pediatric healthcare decision-making. Absence of statistical association between parent and child virtual reality acceptance (χ²(1)=0.11, p=0.74) indicates that virtual reality is effective for both groups independently, enabling implementation approaches that can work with various family situations. These findings collectively indicate that virtual reality interventions can achieve successful multi-stakeholder implementation in pediatric healthcare settings, with consistent satisfaction patterns suggesting robust intervention acceptability for broader clinical implementation across different patient populations and healthcare settings.

### Qualitative Insights and Implementation Lessons

Qualitative analysis revealed meaningful parent perspectives highlighting impact of virtual reality interventions on pediatric healthcare experiences. The predominance of “Engagement and Distraction” as most frequently cited theme (52.4% of responses) aligns with established literature demonstrating that virtual reality’s capacity to redirect children’s attention away from medical procedures represents fundamental mechanism underlying its clinical effectiveness. Parent observations such as “He was distracted and didn’t notice when the needle went in” support research findings that immersive virtual reality environments effectively engage cognitive resources, thereby reducing pain perception through neurological gate control mechanisms.

The “Emotional Impact” theme (34.9% of responses) particularly resonated with parents of children with special needs, with one parent noting enhanced relaxation in their child with autism, reflecting virtual reality’s documented benefits across diverse pediatric populations including those with neurodevelopmental conditions. The emergence of “Clinical Utility” (31.7% of responses) suggests that parents recognized virtual reality’s broader healthcare applications beyond their immediate experience, indicating stakeholder appreciation for intervention’s systemic value in pediatric care settings.

The minimal occurrence of “Challenges and Concerns” (4.8% of responses) mirrors implementation studies showing high acceptability rates and low adverse event profiles for pediatric virtual reality interventions. The nine-month implementation project demonstrated that even busy pediatric preoperative unit can integrate virtual reality without disrupting clinical workflow. Several critical implementation lessons emerged from systematic Plan-Do-Study-Act cycle approach. The iterative refinement process revealed that successful virtual reality integration requires flexibility in session duration, with shift from rigid 3-5 minute protocols to patient-directed 5-20 minute sessions resulting in enhanced effectiveness (48% anxiety reduction for ≥10-minute sessions versus 31% for shorter sessions) while maintaining surgical scheduling efficiency.

The expansion from narrow procedural support to comprehensive preoperative anxiety management significantly broadened intervention’s clinical utility, addressing both acute procedural fears and generalized waiting-period anxiety affecting majority of pediatric patients. Successful integration of family-centered care through tablet-based mirroring addressed parental concerns about content exposure while enhancing collaborative care experience, demonstrating that stakeholder engagement strategies are essential for sustainable implementation. Gradual increase in staff comfort observed across cycles indicates that virtual reality adoption follows predictable learning curve, with nursing satisfaction and workflow integration improving as familiarity increases. Achievement of zero equipment failures and consistent 7.2-minute average setup times in final cycle demonstrates operational sustainability and technical reliability in real-world clinical conditions.

### Predictors of Holistic Implementation Success

The identification of baseline fear score as sole significant predictor of implementation success (odds ratio 2.30, 95% confidence interval 1.09-4.84, p=0.029) supports consistent finding that children with higher initial anxiety levels demonstrate greatest capacity for clinically meaningful improvement through virtual reality distraction. This overall implementation outcome model suggests that fear-based distress represents more robust predictor of comprehensive treatment success than demographic factors or general anxiety measures, supporting targeted virtual reality deployment strategies that prioritize children presenting with elevated procedural fears.

The alignment of positive qualitative themes with quantitative predictors indicates that successful VR implementation depends not only on clinical outcomes but also on stakeholder recognition of the intervention’s engagement capabilities and emotional benefits, providing a foundation for sustainable adoption in pediatric healthcare environments.

### Study Strengths and Limitations

Several strengths enhance confidence in findings and distinguish this investigation from previous research in pediatric virtual reality applications. The real-world clinical implementation represents fundamental strength, as data were collected during actual preoperative care in high-volume pediatric surgical environment rather than controlled research conditions. This real-world validity ensures that findings reflect achievable outcomes under routine clinical practice, accounting for variability, time constraints, and operational complexities of hospital settings.

The iterative Plan-Do-Study-Act methodology used during quality improvement phase enabled continuous refinement and validation of intervention protocols across five systematic cycles. By time data collection was complete, intervention protocol had been pressure-tested and optimized, providing reliability to observed outcomes. This dynamic optimization process, rarely reported in traditional research studies, resulted in evidence-driven modifications such as flexible session durations and parent viewing capabilities that likely contributed to robust outcomes observed in later implementation cycles.

Comprehensiveness of outcome measures represents another notable strength. By including psychological, physiological, and experiential measures, the study provided overall evaluation of intervention impact. Consistency of evidence across reductions in subjective fear and distress, objective physiological measures, and stakeholder satisfaction ratings creates clear narrative of virtual reality’s benefits and minimizes likelihood that results were due to any single type of bias or measurement artifact. Integration of qualitative thematic analysis from parent feedback provides rich contextual understanding of intervention mechanisms, with themes such as “Engagement and Distraction” and “Emotional Impact” directly validating theoretical frameworks underlying virtual reality’s effectiveness as attention-based therapeutic tool.

The custom-designed virtual reality content, developed specifically for target population and clinical setting, enhances internal validity by ensuring that observed benefits can be attributed to purposefully designed virtual reality experience rather than generic technology applications.

Technical reliability demonstrated through zero equipment failures across 80 implementations indicates that current virtual reality hardware meets clinical usage requirements, eliminating technological confounds from outcome assessment and supporting feasibility of virtual reality integration in busy clinical environments. The multidisciplinary collaborative approach involving pediatric physicians, child life specialists, nurses, and research team in both implementing and evaluating virtual reality intervention facilitated systematic refinement through quality improvement cycles while ensuring findings are credible and practical for similar pediatric healthcare settings.

Despite these contributions, several methodological limitations must be acknowledged. The single-arm, pre-post study design without control group represents most significant limitation, preventing definitive causal attribution of observed improvements solely to virtual reality intervention. Natural anxiety reduction over time, environmental acclimation, or placebo effects could potentially account for some observed changes, although magnitude and consistency of improvements combined with stakeholder observations suggest that temporal factors alone cannot explain observed outcomes. Potential for response bias constitutes another limitation, as participants’ awareness of receiving novel intervention may have generated positive expectations that influenced self-reported outcomes through demand characteristics or social desirability bias. Generalizability limitations arise from single-center academic medical center setting and sample characteristics, including predominantly educated parents with previous healthcare experience and children aged 6-15 years undergoing elective procedures. These factors may limit applicability to community hospitals, emergency surgical contexts, or populations with different technological familiarity or healthcare literacy levels. Measurement limitations include reliance on brief, simplified anxiety scales that, while practical and child-friendly, may lack sensitivity of comprehensive anxiety assessments and could contribute to ceiling or floor effects. The immediate-term outcome assessment represents another limitation, as longitudinal follow-up was not conducted to evaluate whether preoperative anxiety reduction translated to improved anesthesia induction compliance, reduced postoperative complications, or enhanced recovery outcomes. The composite “implementation success” measure, while conceptually useful, was not formally validated and should be interpreted cautiously given its modest explanatory power (pseudo R²=0.12).

Finally, inability to blind participants or providers to virtual reality intervention could introduce bias in outcome reporting or patient management, although multi-stakeholder consistency of positive findings and objective physiological improvements support validity of observed effects. Despite these limitations, multi-faceted consistency of results, methodological rigor of implementation approach, and comprehensive stakeholder evaluation provide credible foundation for supporting virtual reality integration into pediatric preoperative care protocols while identifying clear directions for future randomized controlled investigations.

### Clinical and Research Implications

This study provides preliminary support for incorporating VR as a routine, non-pharmacological intervention for managing pediatric preoperative anxiety. The level of anxiety reduction observed—particularly among children with high baseline distress—represents a clinically meaningful improvement that complements existing approaches. Successful integration will require institutional investment in equipment, staff training, and workflow adaptation, but our findings show that VR can be implemented in busy surgical environments without disrupting care. Iterative refinements, including flexible session durations and parent engagement strategies, enhanced both effectiveness and acceptability, offering a practical framework for replication in other pediatric healthcare systems.

Importantly, results highlight the value of patient-tailored use. Children with high baseline fear derived the greatest benefit, suggesting that preoperative screening could guide prioritization when resources are limited. Nevertheless, given the high satisfaction and absence of adverse effects, offering VR broadly remains reasonable, with targeted prioritization improving efficiency in resource-constrained settings.

Future research should focus on rigorous randomized controlled trials comparing VR with both standard care and active non-pharmacological comparators. These studies should include economic evaluations to assess cost-effectiveness relative to pharmacological premedication and workflow efficiency. Multi-center implementation studies are also needed to test scalability across diverse healthcare environments. Additional research should explore optimization of VR content (e.g., immersive environments vs. interactive platforms), timing of delivery (anticipatory preparation vs. intraoperative distraction), and potential family-centered applications, including parent-directed VR protocols.

Finally, implementation science investigations should address practical challenges of scaling VR programs, including training models, workflow integration, and quality assurance mechanisms. Together, these directions will support the transition of VR from a promising innovation to an evidence-based standard of care in pediatric perioperative practice.

## Conclusions

This study demonstrates that carefully implemented virtual reality (VR) can meaningfully improve the preoperative experience for children and their families. VR significantly reduced distress, fear, and physiological indicators of anxiety, while achieving high acceptance among children, parents, and healthcare providers. By evaluating psychological, physiological, and experiential outcomes, we showed that VR is not only clinically effective but also feasible for integration into routine pediatric workflows.

The innovative use of custom pediatric-specific VR content, combined with an iterative quality improvement process, provides a practical framework for broader adoption. Importantly, findings suggest that children with the greatest baseline anxiety may experience the largest benefits, highlighting the potential value of targeted deployment strategies.

For healthcare systems, these results indicate that investing in VR could enhance patient experience, reduce anxiety-related complications, and support staff workflow. For researchers, the work offers implementation guidance and underscores the need for rigorous trials to confirm efficacy, explore long-term outcomes, and evaluate cost-effectiveness.

With continued research and thoughtful implementation, VR has the potential to become a standard component of compassionate, family-centered pediatric care—helping more children approach medical procedures with confidence and calm.

## Data Availability

Upon acceptance for publication, all data will be made available either via a github repository

## ACKNOWLEDGMENTS

We thank the preoperative team at Children’s Hospital of Richmond at VCU for supporting the quality improvement initiative. We acknowledge Suad Alshammari, Ali Alsuhibani, Silas Contaifer, and Ali Alghubayshi for their assistance with data collection. We are grateful to the families who participated and provided feedback.

